# Cost-effectiveness of implementing objective diagnostic verification of asthma in the United States

**DOI:** 10.1101/19012435

**Authors:** Mohsen Yaghoubi, Amin Adibi, Zafar Zafari, J Mark FitzGerald, Shawn D. Aaron, Kate M. Johnson, Mohsen Sadatsafavi

## Abstract

**Background:** Asthma diagnosis in the community is often made without objective testing.

**Objective:** The aim of this study was to evaluate the cost-effectiveness of implementing a stepwise objective diagnostic verification algorithm among patients with community-diagnosed asthma in the United States (US).

**Methods:** We developed a probabilistic time-in-state cohort model that compared a stepwise asthma verification algorithm based on spirometry and methacholine challenge test against the current standard of care over 20 years. Model input parameters were informed from the literature and with original data analyses when required. The target population was US adults (≥15 y/o) with physician-diagnosed asthma. The final outcomes were costs (in 2018 $) and quality-adjusted life years (QALYs), discounted at 3% annually. Deterministic and probabilistic analyses were undertaken to examine the effect of alternative assumptions and uncertainty in model parameters on the results.

**Results:** In a simulated cohort of 10,000 adults with diagnosed asthma, the stepwise algorithm resulted in the removal of diagnosis in 3,366. This was projected to be associated with savings of $36.26 million in direct costs and a gain of 4,049.28 QALYs over 20 years. Extrapolating these results to the US population indicated an undiscounted potential savings of $56.48 billion over 20 years. Results were robust against alternative assumptions and plausible changes in values of input parameters.

**Conclusion:** Implementation of a simple diagnostic testing algorithm to verify asthma diagnosis might result in substantial savings and improvement in patients’ quality of life.

**Key Messages:**

- Compared with current standards of practice, the implementation of an asthma verification algorithm among US adults with diagnosed asthma can be associated with reduction in costs and gain in quality of life.
- There is substantial room for improving patient care and outcomes through promoting objective asthma diagnosis.

**Capsule summary:** Asthma ‘overdiagnosis’ is common among US adults. An objective, stepwise verification algorithm for re-evaluation of asthma diagnosis can result in substantial savings in costs and improvements in quality of life.

## Introduction

Asthma is a common chronic respiratory disease, globally and in the United States (US)^1^. According to the US Centers for Disease Control and Prevention, asthma prevalence continues to rise; the age-standardized prevalence increased by 1.5% per year since 2001, reaching 8.4% in 2010^2^. There are approximately 11 million patients with physician-diagnosed asthma in the US^3^, and total costs of asthma in the US was estimated to be $81.9 billion in 2013^4^. The annual incremental direct medical costs of adult asthma were reported to be $3,266 per patient per year (in 2016 $)^4^.

Evidence-based guidelines recommend that the diagnosis of asthma be made based on objective testing for reversible airflow obstruction or variability in lung function^5^. In some patients spirometry testing may be inconclusive; such patients may require provocative testing to document the presence of airway hyper-responsiveness. In reality, for many Americans, the diagnosis of asthma is made on clinical grounds alone^6,7^. This approach can be inaccurate and can result in labelling individuals with asthma who otherwise have occasional respiratory symptoms without airflow obstruction. In addition, many asthma patients experience spontaneous clinical remission^8^. While the underlying inflammatory process can remain active, such patients may not experience symptoms of asthma for long periods of time even if they discontinue their therapies^9–11^. Due to these factors, a significant proportion of asthma patients in the community might be inappropriately treated. In a multicenter, population-based prospective Canadian cohort study, Aaron et al followed 613 participants who reported a physician diagnosis of asthma within the previous five years and applied an objective algorithm to verify the existing asthma diagnosis. They found that more than 30% of participants could have their current diagnosis of asthma ruled out and safely discontinue their medications^10^.

Extrapolating these figures to the US indicates that there are potentially more than 3 million adults with an over-diagnosis of asthma. As such, the economic burden of over-diagnosed asthma warrants attention. Medication costs are the largest component of the direct economic burden of asthma, contributing to over half of the disease-related costs^4^. Patients with misdiagnosed asthma will incur these costs without gaining benefit, and are subject to harm due to the adverse effects of therapy as well as the lost opportunity to appropriately treat other health conditions that might be responsible for their symptoms.

In this context, a pressing public health question is whether a secondary verification of asthma diagnosis can be considered an efficient use of healthcare resources. A previous study on the economic impact of asthma screening strategies was limited to assessing asthma-related cost savings from implementing a secondary screening program in Canada^12^. This study did not evaluate the effects on health outcomes, such as asthma control and quality of life.

The purpose of the current study was to evaluate the cost-effectiveness of objective diagnostic testing for asthma among previously diagnosed adults in the US. We calculated the incremental average costs and quality-adjusted life years (QALYs) associated with the implementation of objective diagnostic testing. We extrapolated the results to the entire US adult asthma population to evaluate the population-level impact of implementing such a strategy.

## Methods

In reporting on the study’s methodology and results we followed the Consolidated Health Economic Evaluation Reporting Standards^13^. The target population for this cost-effectiveness analysis was US adults (≥15 y/o) with self-reported physician-diagnosed asthma. The diagnostic verification was assumed to be conducted during routine outpatient encounters (e.g., during a scheduled follow-up visit). The main outcomes of the analysis were the incremental costs and QALYs associated with the use of diagnostic verification, compared with the current standard of care *(status quo:* no further verification of diagnosed asthma). We adopted a third-party payer’s perspective (e.g., a Health Maintenance Organization) in the main analysis, and a societal perspective in a secondary analysis. The time horizon was 20 years, and costs and QALYs were discounted at the rate of 3% based on recommendations of the US Panel on Cost Effectiveness in Health and Medicine^14^.

### The intervention

The comparator, the *status quo*, was the continuation of asthma management without any further evaluation of diagnosis. The intervention was defined as a two-stage diagnostic verification algorithm, as illustrated in ***Figure 1***. In accordance with care standards set by the Global Initiative for Asthma (GINA)^15^, the first step is spirometry with reversibility testing before and after inhaled bronchodilation. Asthma is verified in this step if the forced expiratory volume in the first second of expiration (FEV_1_) improves by at least 12% and by 200 millilitres^15^. Those who do not exhibit reversible airflow obstruction undergo a single methacholine challenge test (MCT) at a second visit. Individuals whose FEV_1_ decreases by 20% or more after breathing up to 8 mg/ml of methacholine (in a stepwise fashion based on doubling the methacholine dose) are considered to have their asthma confirmed. Individuals whose MCT is negative are considered to have their asthma ruled out and it was assumed that their asthma-related medications could be tapered off. The use of one MCT, as opposed to up to three MCTs implemented in the research context, was considered to be the most realistic scenario for clinical practice.

**Figure 1:**
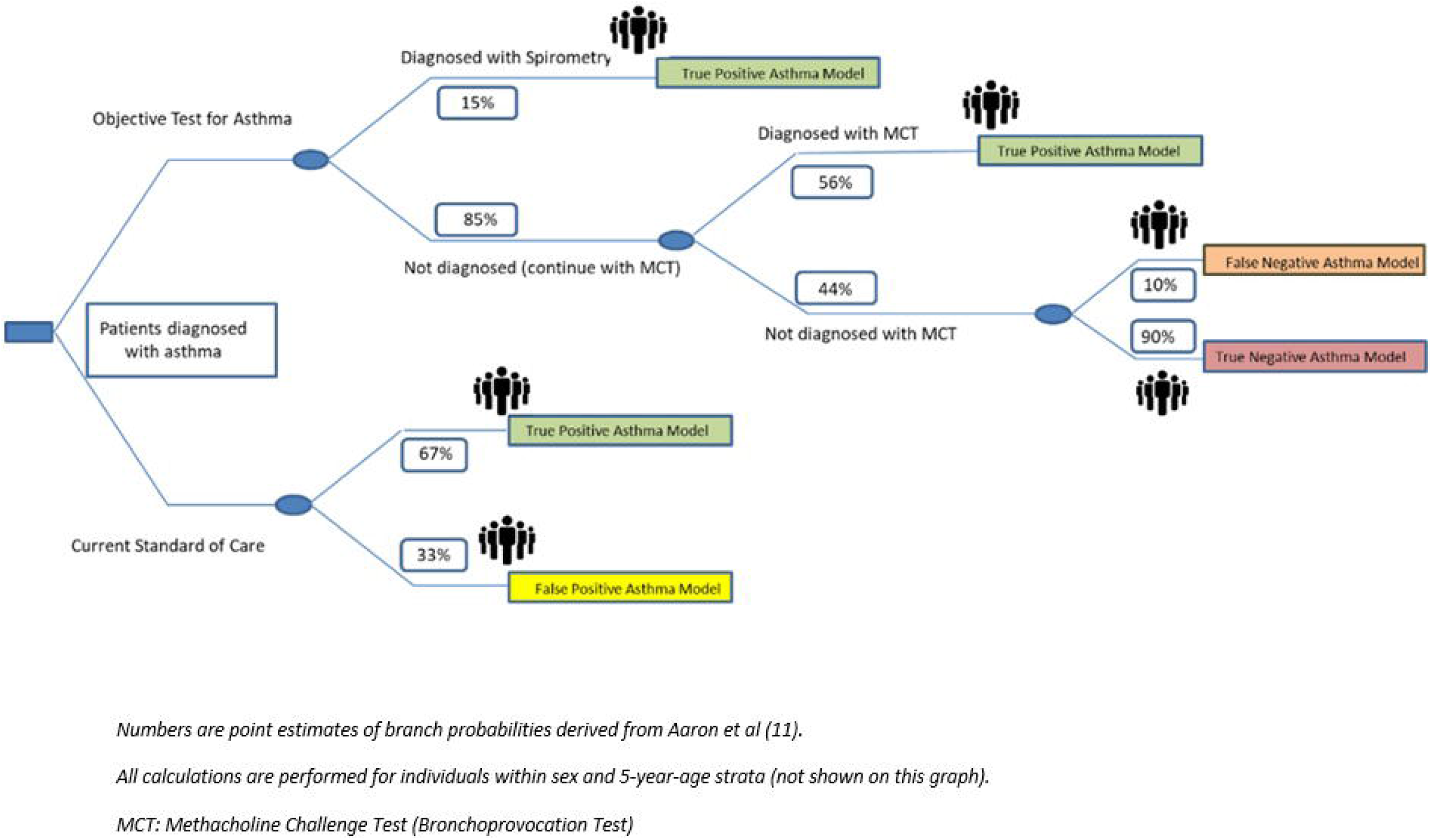
Algorithm for stepwise asthma verification testing.

### Modeling approach

We constructed a decision tree to model the diagnostic steps, and a time-in-state model to follow individuals at terminal branches of the tree over the analysis’ time horizon in annual cycles. The model stratified US adult (≥15 years old) with diagnosed asthma into age (with 5-year intervals) and sex groups. A schematic illustration of the decision tree, with point estimates of branch probabilities, is provided in ***Figure 2***. The major input parameters for the model are provided in ***Table 1***.

**Table 1).**
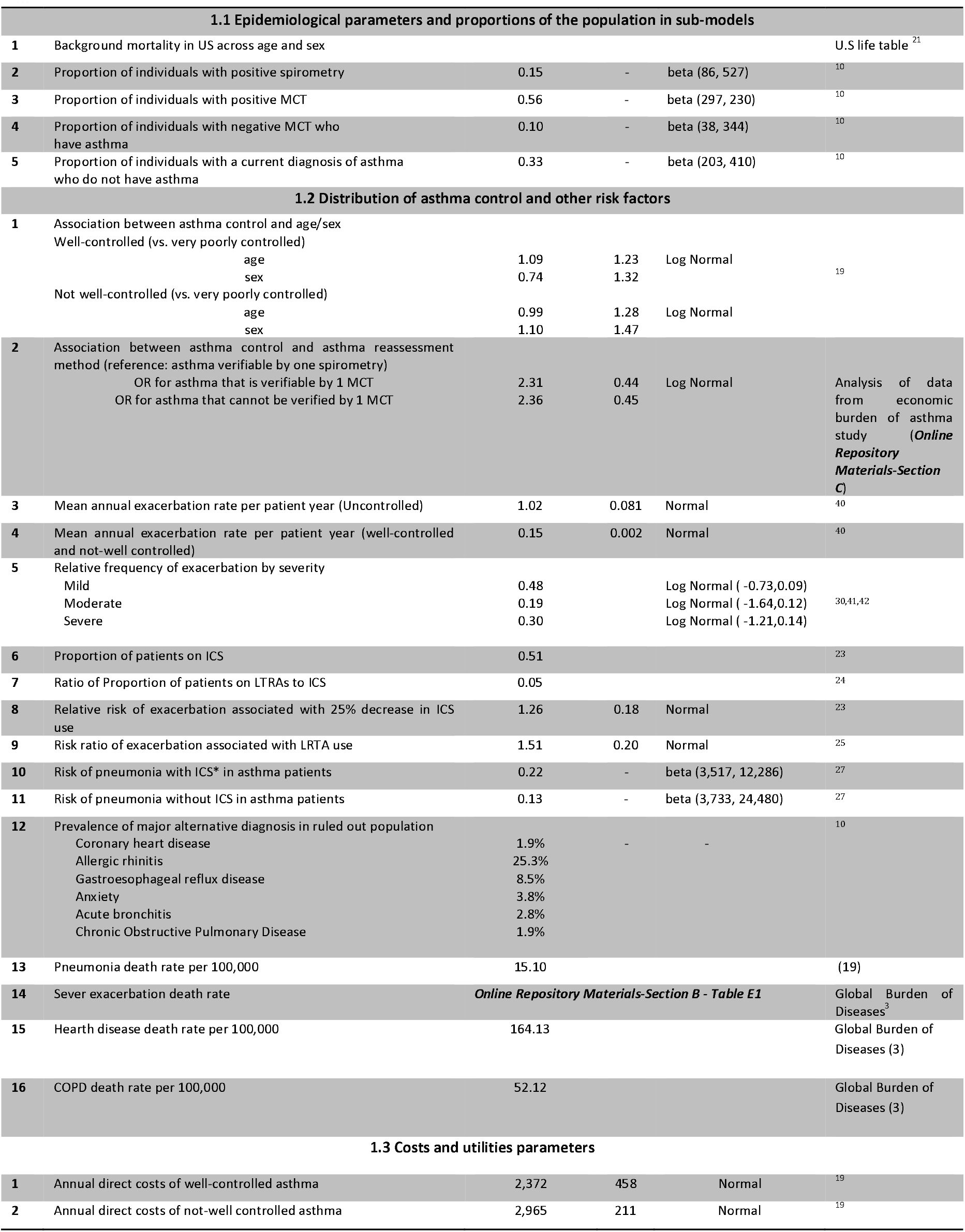
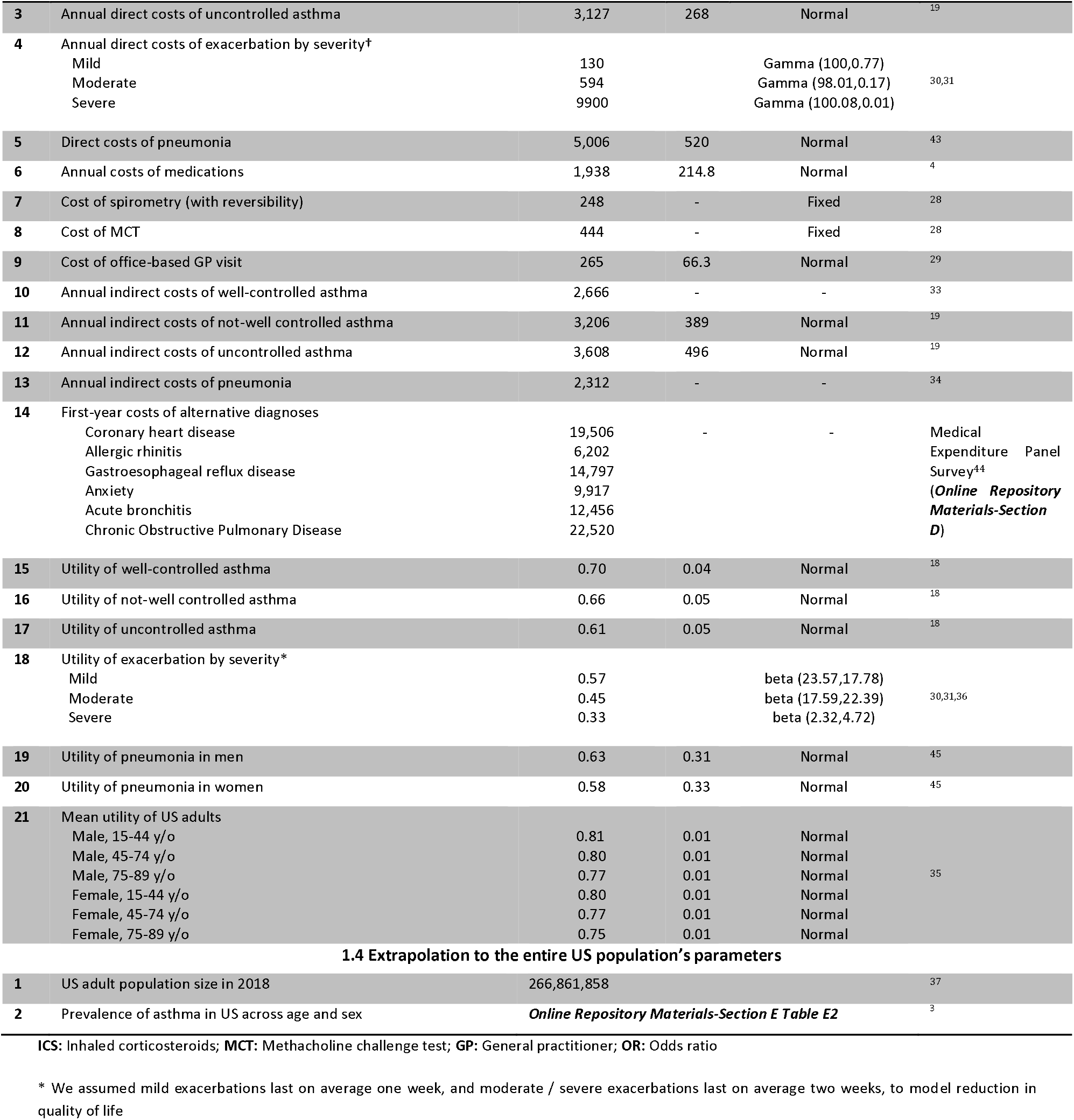
Input parameters.

| 1.1 Epidemiological parameters and proportions of the population in sub-models |  |  |  |  |  |
| --- | --- | --- | --- | --- | --- |
| 1 | Background mortality in US across age and sex |  |  |  | U.S life table <sup>21</sup> |
| 2 | Proportion of individuals with positive spirometry | 0.15 | - | beta (86, 527) | 10 |
| 3 | Proportion of individuals with positive MCT | 0.56 | - | beta (297, 230) | 10 |
| 4 | Proportion of individuals with negative MCT who have asthma | 0.10 | - | beta (38, 344) | 10 |
| 5 | Proportion of individuals with a current diagnosis of asthma who do not have asthma | 0.33 | - | beta (203, 410) | 10 |
| 1.2 Distribution of asthma control and other risk factors |  |  |  |  |  |
| 1 | Association between asthma control and age/sex |  |  |  |  |
|  | Well-controlled (vs. very poorly controlled) |  |  |  |  |
|  | age | 1.09 | 1.23 | Log Normal | 19 |
|  | sex | 0.74 | 1.32 |  |  |
|  | Not well-controlled (vs. very poorly controlled) |  |  |  |  |
|  | age | 0.99 | 1.28 | Log Normal |  |
|  | sex | 1.10 | 1.47 |  |  |
| 2 | Association between asthma control and asthma reassessment method (reference: asthma verifiable by one spirometry) |  |  |  |  |
|  | OR for asthma that is verifiable by 1 MCT | 2.31 | 0.44 | Log Normal | Analysis of data from economic burden of asthma study (*Online Repository Materials-Section C*) |
|  | OR for asthma that cannot be verified by 1 MCT | 2.36 | 0.45 |  |  |
| 3 | Mean annual exacerbation rate per patient year (Uncontrolled) | 1.02 | 0.081 | Normal | 40 |
| 4 | Mean annual exacerbation rate per patient year (well-controlled and not-well controlled) | 0.15 | 0.002 | Normal | 40 |
| 5 | Relative frequency of exacerbation by severity |  |  |  |  |
|  | Mild | 0.48 |  | Log Normal ( -0.73,0.09) | 30,41,42 |
|  | Moderate | 0.19 |  | Log Normal ( -1.64,0.12) |  |
|  | Severe | 0.30 |  | Log Normal ( -1.21,0.14) |  |
| 6 | Proportion of patients on ICS | 0.51 |  |  | 23 |
| 7 | Ratio of Proportion of patients on LTRAs to ICS | 0.05 |  |  | 24 |
| 8 | Relative risk of exacerbation associated with 25% decrease in ICS use | 1.26 | 0.18 | Normal | 23 |
| 9 | Risk ratio of exacerbation associated with LRTA use | 1.51 | 0.20 | Normal | 25 |
| 10 | Risk of pneumonia with ICS* in asthma patients | 0.22 | - | beta (3,517, 12,286) | 27 |
| 11 | Risk of pneumonia without ICS in asthma patients | 0.13 | - | beta (3,733, 24,480) | 27 |
| 12 | Prevalence of major alternative diagnosis in ruled out population |  |  |  | 10 |
|  | Coronary heart disease | 1.9% | - | - |  |
|  | Allergic rhinitis | 25.3% |  |  |  |
|  | Gastroesophageal reflux disease | 8.5% |  |  |  |
|  | Anxiety | 3.8% |  |  |  |
|  | Acute bronchitis | 2.8% |  |  |  |
|  | Chronic Obstructive Pulmonary Disease | 1.9% |  |  |  |
| 13 | Pneumonia death rate per 100,000 | 15.10 |  |  | (19) |
| 14 | Sever exacerbation death rate | *Online Repository Materials-Section B - Table E1* |  |  | Global Burden of Diseases <sup>3</sup> |
| 15 | Hearth disease death rate per 100,000 | 164.13 |  |  | Global Burden of Diseases (3) |
| 16 | COPD death rate per 100,000 | 52.12 |  |  | Global Burden of Diseases (3) |
| 1.3 Costs and utilities parameters |  |  |  |  |  |
| 1 | Annual direct costs of well-controlled asthma | 2,372 | 458 | Normal | 19 |
| 2 | Annual direct costs of not-well controlled asthma | 2,965 | 211 | Normal | 19 |
| 3 | Annual direct costs of uncontrolled asthma | 3,127 | 268 | Normal | 19 |
| 4 | Annual direct costs of exacerbation by severity† |  |  |  |  |
|  | Mild | 130 |  | Gamma (100,0.77) |  |
|  | Moderate | 594 |  | Gamma (98.01,0.17) | 30,31 |
|  | Severe | 9900 |  | Gamma (100.08,0.01) |  |
| 5 | Direct costs of pneumonia | 5,006 | 520 | Normal | 43 |
| 6 | Annual costs of medications | 1,938 | 214.8 | Normal | 4 |
| 7 | Cost of spirometry (with reversibility) | 248 | - | Fixed | 28 |
| 8 | Cost of MCT | 444 | - | Fixed | 28 |
| 9 | Cost of office-based GP visit | 265 | 66.3 | Normal | 29 |
| 10 | Annual indirect costs of well-controlled asthma | 2,666 | - | - | 33 |
| 11 | Annual indirect costs of not-well controlled asthma | 3,206 | 389 | Normal | 19 |
| 12 | Annual indirect costs of uncontrolled asthma | 3,608 | 496 | Normal | 19 |
| 13 | Annual indirect costs of pneumonia | 2,312 | - | - | 34 |
| 14 | First-year costs of alternative diagnoses |  |  |  |  |
|  | Coronary heart disease | 19,506 | - | - | Medical |
|  | Allergic rhinitis | 6,202 |  |  | Expenditure Panel |
|  | Gastroesophageal reflux disease | 14,797 |  |  | Survey<sup>44</sup> |
|  | Anxiety | 9,917 |  |  | (Online Repository |
|  | Acute bronchitis | 12,456 |  |  | Materials-Section |
|  | Chronic Obstructive Pulmonary Disease | 22,520 |  |  | D) |
| 15 | Utility of well-controlled asthma | 0.70 | 0.04 | Normal | 18 |
| 16 | Utility of not-well controlled asthma | 0.66 | 0.05 | Normal | 18 |
| 17 | Utility of uncontrolled asthma | 0.61 | 0.05 | Normal | 18 |
| 18 | Utility of exacerbation by severity* |  |  |  |  |
|  | Mild | 0.57 |  | beta (23.57,17.78) |  |
|  | Moderate | 0.45 |  | beta (17.59,22.39) | 30,31,36 |
|  | Severe | 0.33 |  | beta (2.32,4.72) |  |
| 19 | Utility of pneumonia in men | 0.63 | 0.31 | Normal | 45 |
| 20 | Utility of pneumonia in women | 0.58 | 0.33 | Normal | 45 |
| 21 | Mean utility of US adults |  |  |  |  |
|  | Male, 15-44 y/o | 0.81 | 0.01 | Normal |  |
|  | Male, 45-74 y/o | 0.80 | 0.01 | Normal |  |
|  | Male, 75-89 y/o | 0.77 | 0.01 | Normal | 35 |
|  | Female, 15-44 y/o | 0.80 | 0.01 | Normal |  |
|  | Female, 45-74 y/o | 0.77 | 0.01 | Normal |  |
|  | Female, 75-89 y/o | 0.75 | 0.01 | Normal |  |
| 1.4 Extrapolation to the entire US population's parameters |  |  |  |  |  |
| 1 | US adult population size in 2018 | 266,861,858 |  |  | 37 |
| 2 | Prevalence of asthma in US across age and sex | Online Repository Materials-Section E Table E2 |  |  | 3 |

ICS: Inhaled corticosteroids; MCT: Methacholine challenge test; GP: General practitioner; OR: Odds ratio
\* We assumed mild exacerbations last on average one week, and moderate / severe exacerbations last on average two weeks, to model reduction in quality of life

**Figure 2:**
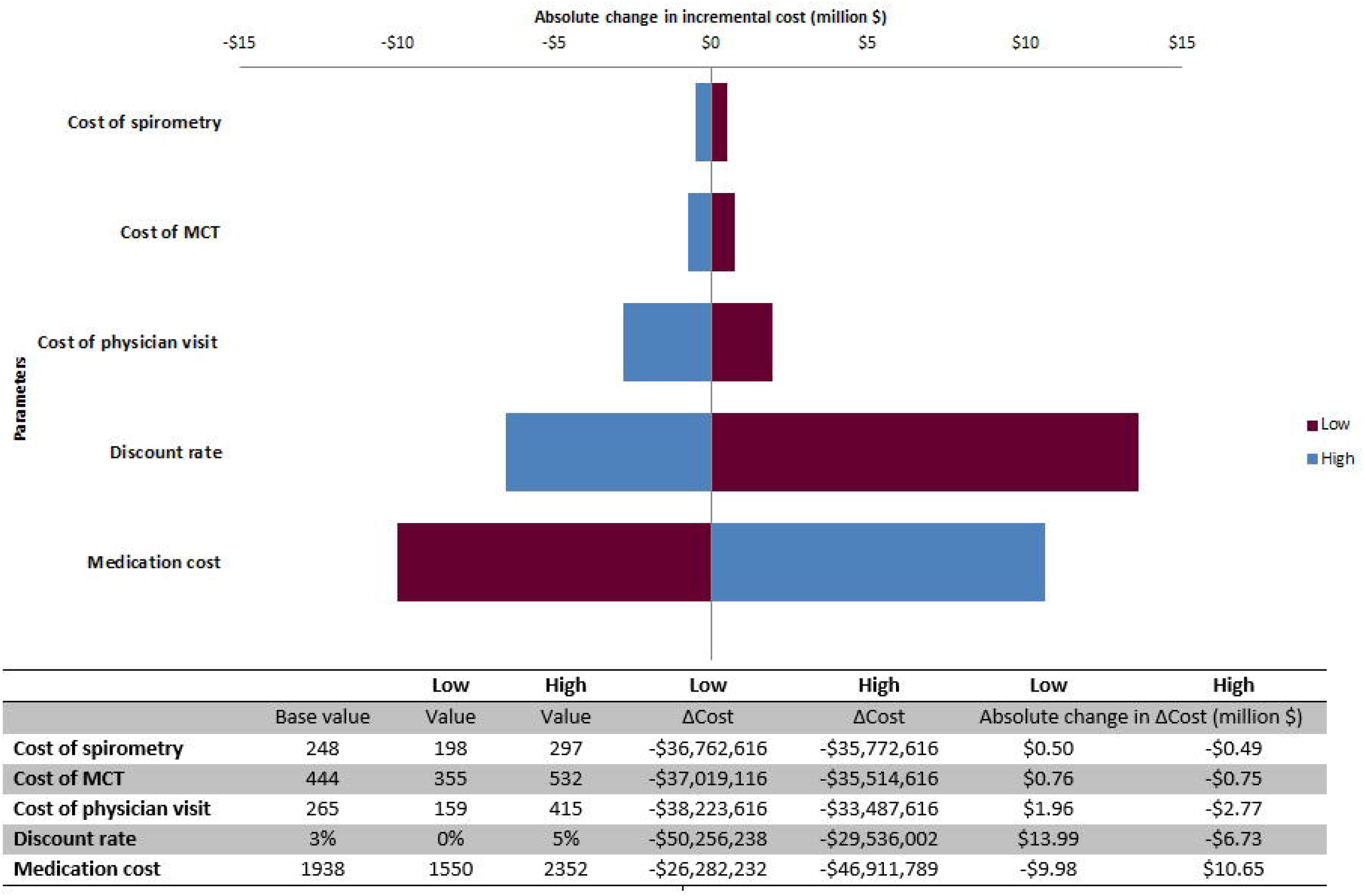
Decision tree combined with open population time-in-state model. Model used for evaluating costs and health-related outcomes of asthma verification national program compared to status quo.

The proportion of individuals at each branch of the decision tree was directly taken from the population-based study by Aaron et al^10^. In the *status quo* scenario, 33% of individuals were assumed not to have asthma. These are the individuals who responded negatively to spirometry and three subsequent MCTs in the above-mentioned study. Given that there will be no verification of diagnosis, these individuals remained ‘over-diagnosed’ throughout the time horizon of the study (and as such continued to be treated for asthma) under the *status quo* strategy.

In the diagnostic verification scenario, 15% of patients will have their asthma confirmed through a positive spirometry; of the remaining, 48% will have their asthma confirmed through a positive MCT. A diagnosis of asthma in individuals with a negative spirometry and a negative MCT will be considered to have been ruled out and these individuals will be advised to taper their medications. In the study by Aaron et al, in 10% of such individuals the asthma diagnosis was later confirmed with a positive second or third MCT, after tapering of controller medications (***Table 1*** – section 1.1). This figure is similar to the study by Luks et al ^16^(9%) and reflects imperfect sensitivity of MCT especially when the patient is receiving asthma medications^16,17^. Given that only one MCT is modeled in the diagnostic verification scenario in the present study, such individuals constitute the ‘false negative’ subgroup (those in whom an asthma diagnosis was erroneously reversed), with the rest of individuals under this branch of the decision tree constituting the ‘true negative’ subgroup (***Figure 2***). Compared to their counterparts in the *status quo* scenario, individuals in the false negative asthma subgroup will cease their controller medication, and as such will generally have worse asthma control.

### Modeling asthma status over time

At terminal branches of the decision tree, each of these sub-populations was linked to its corresponding time-in-state model. Such models simulated the transition of a sex- and age-stratified population across mutually exclusive health states. For individuals with either true asthma or misdiagnosed asthma (true positive and false negative subgroups in ***Figure 2***), the model contained three states pertaining to the three levels of asthma control were assigned, in addition to a state representing death. The distribution of levels of asthma control within a given sex and age group was derived using calibration techniques from a recent study based on US National Health and Wellness Survey, as described in our previous work^18,19^ (see distribution of asthma control in ***Online Repository Materials - Section A***). The levels of asthma control were defined based on the asthma control test (ACT). This test classifies a patient’s asthma status into “poorly controlled” (scores ≤ 15), “not well controlled” (score 16-19), and “well controlled” (score 20-25)^20^. In addition to asthma control, we modeled asthma exacerbations for individuals with true asthma as an event that are likely to be affected by controller therapy (see modeling asthma exacerbation in ***Online Repository Materials - Section B***). On the other hand, in the subgroup with true negative and false positive asthma, the model consisted of only two states of “alive” and “dead”. Aside from mortality due to severe exacerbations, the probability of transitioning to death for all populations was equal to the age- and sex-specific background mortality rates, derived from the US life tables^21^.

It is likely that the degree of airway reactivity is an independent determinant of asthma control. As such, the distribution of asthma control levels can be different among individuals whose asthma can be confirmed using spirometry, those whose asthma verification requires an MCT, and those who have asthma but do not display reversibility or have a positive MCT. To elucidate such potential differences, we used data from the Economic Burden of Asthma (EBA) study^22^. This longitudinal study used population-based sampling and followed 622 adult individuals with physician-diagnosed asthma over one year. Asthma status was verified with one pre- and post-bronchodilator spirometry and one MCT (similar to the protocol evaluated in this study). Asthma control was verified during baseline and each of the three-monthly visits. Using these data, we constructed a generalized linear model (with multinomial distribution and cumulative logit link) to estimate the odds ratio (OR) of having controlled asthma compared with uncontrolled asthma, as well as having partially controlled asthma compared with uncontrolled asthma, among the three-aforementioned groups. The exposure was asthma verification status (response to spirometry, response to MCT, no response to spirometry and MCT). The model was adjusted for sex, age at baseline, and whether the individual used any controller medication in the 12 months before the study began (see ***Online Repository Materials-Section C*** and ***Table 1*** – section 1.2 for distribution of asthma control).

### Modeling asthma treatment

A major benefit of the objective diagnostic verification would be the removal of asthma therapies in patients who do not need them. We modeled the utilization and short-term effects of inhaled corticosteroids (ICS) and leukotriene receptor antagonists (LTRA), namely in terms of improving asthma control and reducing exacerbation rates. We assumed that 51% of diagnosed asthma patients are on maintenance ICS therapy at any given time^23^, while an additional 5% are on LTRAs^24^. The use of reliever (rescue) medication was not directly modeled as their costs are embedded within the costs assigned to each control level, and it was assumed their usage will not have any direct impact on asthma control. To model the association between the use of controller medications and asthma control, we used the indirect evidence on the effect of ICS or LTRA use and asthma exacerbations, as performed in our previous modeling study^23,25,26^. We did not include the longer-term adverse events associated with controller medications, but in a sensitivity analysis modeled the increased risk of acute pneumonia as an adverse effect of ICS^27^.

#### Costs

Details on the cost parameters are provided in ***Table 1*** – section 1.3. In the base case analysis, costs included the cost of stepwise asthma verification testing, controller medication costs, direct asthma-attributable costs across levels of control, and direct medical costs resulting from asthma exacerbations. The costs of spirometry testing and MCT were derived from American Thoracic Society Coding and Billing Quarterly^28^ for hospital-based lung function testing. In the intervention scenario, we assumed that interpreting the results of spirometry and MCT will each require one additional physician visit. The costs of office**-**based physician visit were derived from the Medical Expenditure Panel Survey (MEPS) based on average expenses per visit, assuming the mixture of general practitioner and specialist visits as observed in the MEPS data^29^. In a sensitivity analysis, we varied this costs across the entire range of speciality types to cover scenarios that are based on general practitioner or specialist management only. Annual per-person direct medical costs of asthma (including healthcare provider visits, emergency visits, hospitalizations, and medication use) across levels of control (not including exacerbation costs or costs of controller use, as they were explicitly modeled) were derived from our previous work^19^. Costs of exacerbations were obtained from the literature^30,31^.

All costs were adjusted to 2018 USD ($) using historical inflation rates^32^. In the base case analysis we excluded indirect costs based on the recommendation of the US Panel on Cost-Effectiveness in Health and Medicine^14^. In a scenario analysis, we adopted a societal perspective and included costs due to asthma-related productivity loss (indirect costs). Indirect costs due to sub-optimal asthma control were derived from multiple sources^19,33,34^. These included costs due to total cessation of work as well as loss of work time. Indirect costs per level of control included loss of productivity due to exacerbations. We did not include in the base case analysis the costs of work-up required for alternative diagnoses when asthma diagnosis was reversed. However, in a sensitivity analysis, we considered the firs-year costs of six common and major alternative diagnoses as reported by Aaron et al^10^. These costs were derived from a dedicated analysis of 2017 MEPS data (details are provided in ***Online Repository Materials - section D***).

#### Health State Utility Values

The health state utility values (utilities) associated with asthma across levels of control were derived from a study conducted on 2011–2013 US National Health and Wellness Survey^18^. We assigned US-specific general population utilities to individuals whose asthma was ruled out^35^. Utilities associated with different severities of exacerbation were derived from previous studies^30,31,36^ (***Table 1*** – section 1.3).

### Analysis

In the base case analysis, we ran the model separately in the intervention and *status quo* scenarios and estimated total discounted costs and QALYs over the 20-year time horizon. We standardized the results such that they pertain to the outcome of a hypothetical cohort of 10,000 representative individuals.

Deterministic sensitivity and scenario analyses were conducted to investigate the impact of key parameter values and assumptions on the base case results. In addition, we performed two threshold analyses, one on the prevalence of asthma overdiagnosis, and another on the first-year diagnostic and management costs of alternative diagnoses. These analyses identified the critical value of their respective parameters around which the overall findings on the cost-effectiveness of the intervention would change. To fully account for uncertainty in all parameters, probabilistic sensitivity analyses were conducted through Monte Carlo simulations. Uncertainty in each of the underlying parameters was characterized by assigning a probability distribution to the parameter, and the model was run 10,000 times; within each run a new random value from each distribution was drawn and results were recalculated. Cost-effectiveness was assessed by using willingness-to-pay (WTP) thresholds of$50,000/QALY.

Furthermore, we extrapolated the result to the entire US asthma population, using the general methodology as described in our recent work estimating the burden of uncontrolled asthma in the US^19^. In summary, projections of US population by state were derived from the Census Bureau - Population Division^37^. The prevalence of physician-diagnosed asthma stratified by age and sex was obtained from the Global Burden of Disease studies^3^ (***Online Repository Materials - Section E, Tables E2 & E3***). The baseline year was 2019 and cumulative projections were made to 2038 (20 years).

## Results

***Table 2*** provides the results of base case analysis for a cohort of 10,000 adults with physician-diagnosed asthma under the diagnostic verification versus *status quo* scenarios. In the diagnostic verification scenario, in 1,500 patients asthma diagnosis will be confirmed through spirometry alone. Among the 8,500 that will undergo a subsequent MCT, asthma diagnosis will be confirmed in 4,760. Asthma will be ruled out in the remaining 3,740 patients in whom spirometry and MCT will be negative. Among these, in 374 the asthma diagnosis will be erroneously reversed. The total costs of spirometry and MCT in the diagnostic verification scenario will be $2,480,000 and $3,774,000, respectively.

**Table 2.** Results of cost-effectiveness analysis.

| Item | Intervention | Status Quo | Incremental |
| --- | --- | --- | --- |
| Total patients | 10,000 | 10,000 | 0 |
| Number of patients confirmed asthma by spirometry | 1,500 | 0 | 1,500 |
| Number of patients confirmed asthma by MCT* | 4,760 | 0 | 4,760 |
| Number of erroneous rule-outs of asthma | 374 | 0 | 374 |
| Number of correct rule-outs of asthma | 3,366 | 0 | 3,366 |
| Total number of spirometry tests required | 10,000 | 0 | 10,000 |
| Total number of MCTs required | 8,500 | 0 | 8,500 |
| Total patient-years across the stratum | 192,944 | 192,944 | 0 |
| Controlled | 79,404 | 83,511 | -4,107 |
| Partially controlled | 23,821 | 23,028 | 792 |
| Uncontrolled | 27,490 | 24,176 | 3,314 |
| Asthma ruled out | 62,229 | 0 | 62,229 |
| False positive asthma on medication | 0 | 62,229 | -62,229 |
| Total number exacerbations by severity | 33,736 | 32,141 | 1,595 |
| Mild | 16,694 | 15,905 | 789 |
| Moderate | 6,608 | 6,296 | 312 |
| Severe | 10,434 | 9,940 | 493 |
| Number of patient-years on asthma medication | 62,906 | 98,402 | -35,495 |
| Total cost of spirometry | \$2,480,000 | 0 | \$2,480,000 |
| Total cost of MCTs | \$3,774,000 | 0 | \$3,774,000 |
| Total cost of GP visit | \$4,902,500 | 0 | \$4,902,500 |
| Total cost of well-controlled asthma | \$144,570,236 | \$152,046,117 | -7,475,881 |
| Total cost of not well-controlled asthma | \$54,192,213 | \$52,388,430 | 1,803,783 |
| **Total cost of poorly controlled asthma** | **\$65,958,334** | **\$58,005,565** | **7,952,769** |
| **Cost of medication in false positive asthma** | **0** | **\$49,850,476** | **\$-49,850,476** |
| **Total cost of exacerbations** | **\$3,187,580** | **\$3,036,891** | **\$150,689** |
| **Mild** | **\$31,937** | **\$30,427** | **\$1,510** |
| **Moderate** | **\$115,525** | **\$110,063** | **\$5,461** |
| **Severe** | **\$3,040,119** | **\$2,896,401** | **\$143,718** |
| **Total costs** | **\$279,064,863** | **\$315,327,479** | **\$-36,262,616** |
| **Total QALYs** | **105,324.89** | **101,275.61** | **4,049.28** |
| **ICER** | **Dominant\*** |  |  |

**MCT:** methacholine challenge tests; **QALY:** Quality-Adjusted Life Years; **ICER:** Incremental Cost-Effectiveness Ratio
\* The diagnostic verification scenario was associated with lower costs and higher QALYs

Total patient-years on asthma controller medications will be 62,906 in the diagnostic verification and 98,402 in the *status quo* scenario. Total savings in medication costs will be $49.85 million. Under the diagnostic verification scenario, there will be an extra 3,314 patient-years with uncontrolled asthma compared with the *status quo*, and as a result there will be 1,595 more exacerbations. Combining these results, for the diagnostic verification scenario, total discounted costs and QALYs will be $279.06 million and 105,324, respectively. The corresponding values for the *status quo* scenario will be $315.32 million and 101,275. As such, the implementation of the objective asthma verification algorithm will be associated with $36.26 million reduction in direct costs and a gain of 4,049 in QALYs, compared with the *status quo* over the next 20 years.

### Sensitivity and scenario analyses

***Figure 3*** depicts change in incremental costs for different analyses. Diagnostic verification remained cost-saving in all one-way sensitivity analyses. ***Table 3*** presents the results of scenario analyses. Again, the overall conclusion remained the same when several alternative assumptions were explored. The threshold analysis indicated that when the proportion of falsely diagnosed to overall asthma was 6% or lower (compared with 33% in the main analysis), the diagnostic verification was no longer cost-saving. As well, the first-year costs of major alternative diagnoses had to be at least 3 times higher than their original estimates for the diagnostic intervention not to be cost-saving. The inclusion of indirect costs only slightly reduced the difference in costs between the two strategies, and diagnostic intervention remained dominant. Results of the probabilistic sensitivity analysis are provided in ***Figure 4***. In >99% of all simulations, the intervention remained dominant.

**Table 3:**
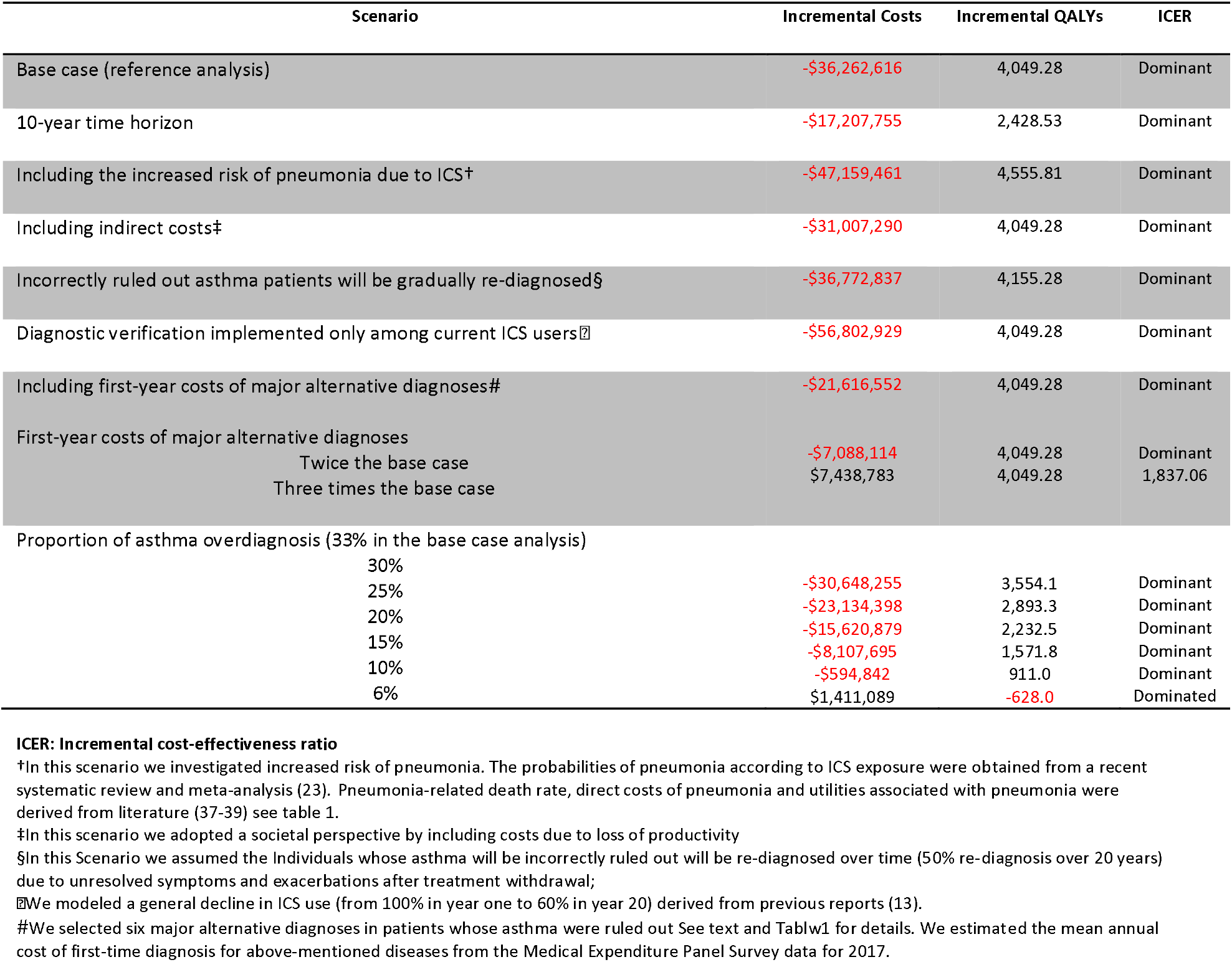
Results of scenario analyses.

**Figure 3.**
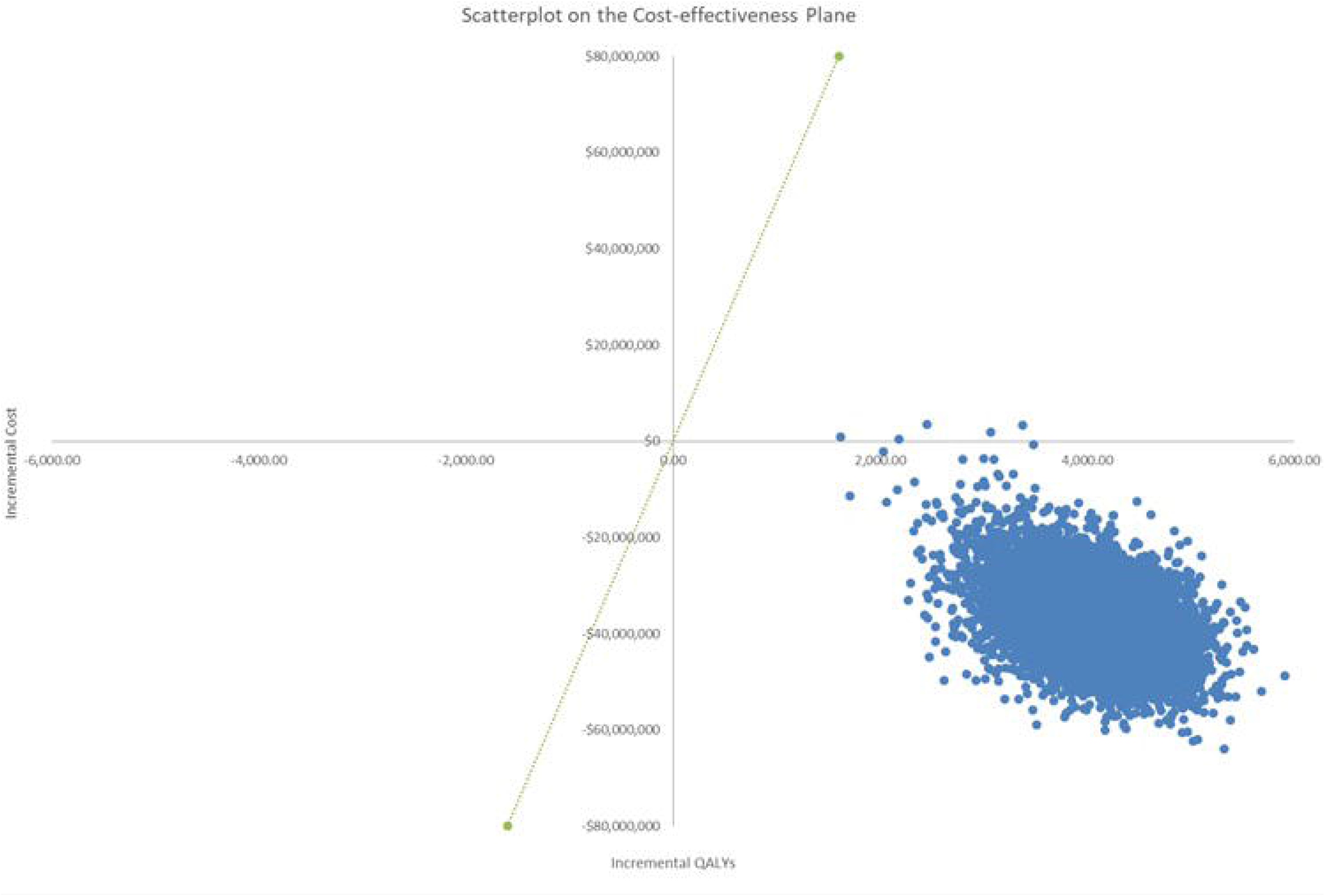
Results of deterministic one-way sensitivity analyses.

**Figure 4:**
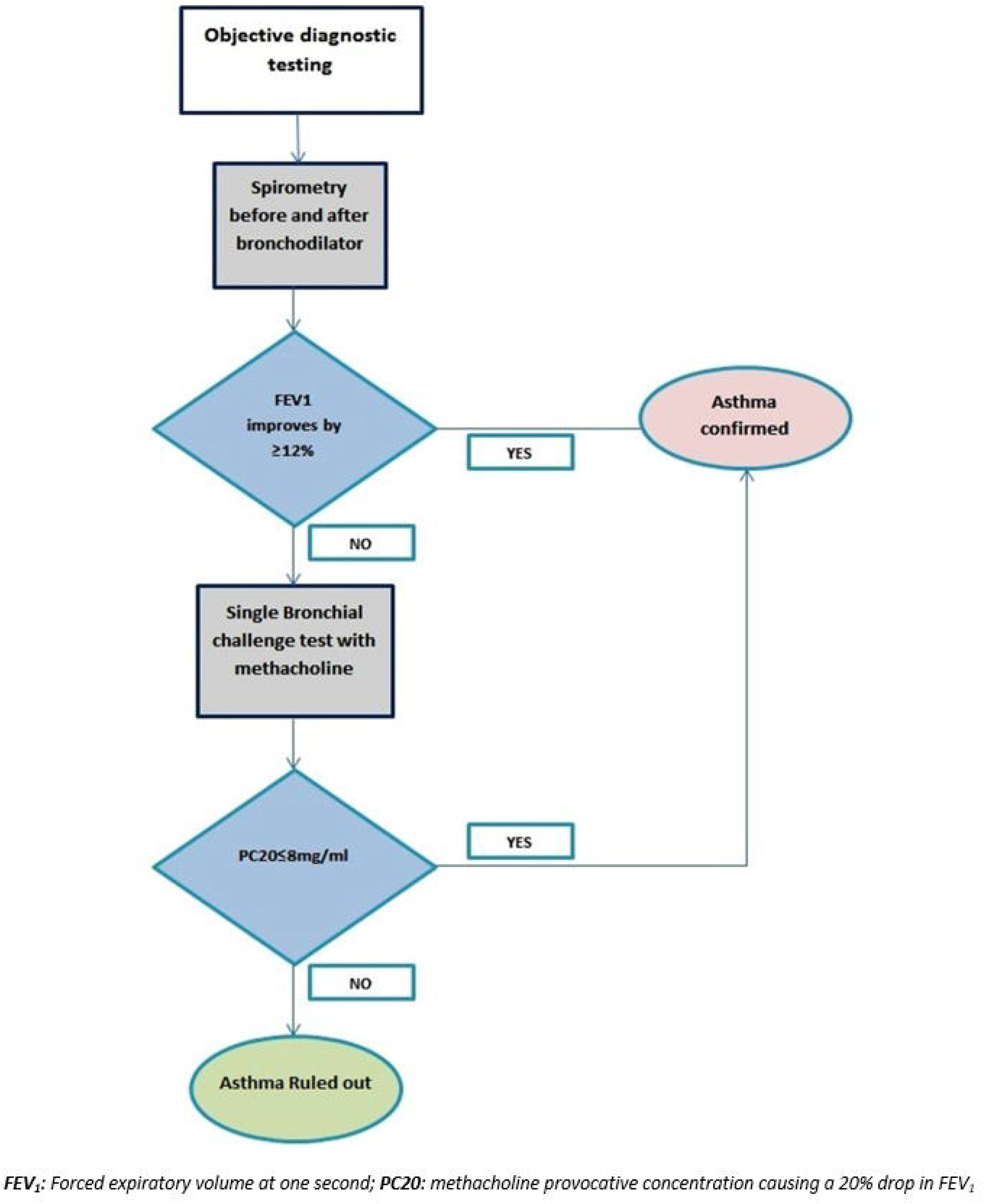
Cost-effectiveness plane for a cohort of 10,000 individuals.

### Extrapolating the results to the entire US population

In 2019, there will be 15.55 million adolescents and adults with asthma in the US; this value will increase by 13% by 2038. After the implementation of diagnostic verification algorithm, during the 2018–2038 period a diagnosis of asthma will be reversed in 70.29 million individuals. The total undiscounted cost saving in the intervention scenario versus *status quo* will be $56.48 billion, and there will be a cumulative gain in QALYs of 6.14 million (see analysis by year for the entire US population in ***Online Repository Materials-Section E and Table E3***).

## Discussion

Our results indicate that the implementation of a stepwise diagnostic verification algorithm at point of care among US adults with a self-reported diagnosis of asthma might be associated with significant savings in costs and improvements in quality of life over twenty years. In the main analysis as well as the sensitivity and scenario analyses, this strategy was dominant against the strategy of continuing asthma management; i.e., it was associated with reduced costs and improvements in quality of life. Extrapolating these results to the entire US population suggested that over 20 years, there could be a saving of $40.51 billion in direct asthma-related medical costs.

These results are despite the fact that our modeling approach was based on several conservative assumptions against diagnostic verification. For one, in patients whose asthma is ruled out, the possible causes of underlying symptoms are more likely to be verified. As such, it is likely that there will be benefits to patients and reductions in costs due to the proper management of the underlying disease. In the study by Aaron et al., more than 50% of patients whose asthma was ruled out were ultimately diagnosed with other conditions. The most common alternative diagnoses were allergic rhinitis, gastro-esophageal reflux disease, anxiety disorders, acute bronchitis, Chronic Obstructive Pulmonary Disease, and Ischemic heart disease. To some extent, the overall efficiency of the asthma verification algorithms is influenced by the efficiency of management of these conditions. It is likely that the additional costs of diagnostic workup and disease management will be offset by better outcomes and increase in quality of life under proper treatment of such conditions. In a sensitivity analysis that explored this aspect, asthma verification test remained cost-saving by incorporating the first-year costs of diagnostic and management of the six most common alternative diagnoses (a finding that remained valid when the prevalence of such conditions were assumed to be up to three times higher than their original values).

Further, we assumed that in around 4 % of individuals asthma will be ruled out incorrectly due to the imperfect sensitivity of spirometry and a single MCT. In such individuals we assumed that controller medications will be discontinued and as result asthma control status will deteriorate. This results in reduced quality of life and a higher rate of exacerbations that will last throughout the time horizon of the study. However, in reality it is likely that the intensification of symptoms and occurrence of exacerbations upon treatment withdrawal will result in further assessments and a re-establishment of the diagnosis. When we modeled this aspect in a sensitivity analysis, the diagnostic intervention scenario was even more cost-saving.

We are aware of one previous economic evaluation of secondary verification of asthma^38^. In the study by Pakhale et al, which was performed in the Canadian context, implementation of an asthma verification algorithm was associated with an average cost savings of $35,141 per 100 screened individuals. The main source of cost saving was medication costs but the authors did not consider the impact of diagnostic verification and subsequent medication changes on asthma control, exacerbations, and quality of life.

The strengths of our study includes its reliance on hierarchy of evidence in estimating model parameters, considering multiple sources of evidence about the burden of uncontrolled asthma and the association between asthma control, treatment, resource use, and quality of life. Nuanced modeling of asthma control over time and how it is affected by diagnostic verification and changes in treatments further enhances the generalizability of our results compared with previous evidence. The incorporation of uncertainty in model inputs and assumptions into the results should provide reassurance that the overall findings are not materially affected by uncertain evidence.

The limitations of our study should also be acknowledged. The main source of evidence on the rate of asthma overdiagnosis was a Canadian study^10^. We could not find any US-based study, but a recent systematic review on this topic reported that studies across different jurisdictions have consistently reported an overdiagnosis rate between 29%-34%^39^. Our threshold analysis also indicated that overdiagnosis rate in the US should be implausibly low for the diagnostic verification not to be cost-saving. We assumed that diagnostic verification will take place in the context of routine clinical practice, as such our results do not pertain to alternative schemes such as a systematic screening program. As well, a strict adherence to the diagnostic verification algorithm might not be achieved in the real world. For example, if the spirometry fails to establish the diagnosis, the physician might decide on a trial of medication cessation and re-evaluation of symptom burden before embarking on the assessment of airway hyperresponsiveness. Such departures from the verification protocol will inevitably affect the cost-effectiveness of diagnostic verification. Furthermore, in extrapolating the results to the entire US, we assumed immediate uptake of the diagnostic verification algorithm while in reality there will be gradual adoption of such a program and imperfect implementation. We did not consider alternative diagnostic strategies such as using the measurement of exhaled nitric oxide as the data for such a diagnostic strategy is less robust. Finally, changes in risk factors as well as future innovations in asthma diagnosis and care can impact the efficiency of the proposed intervention.

There are several opportunities for future research in this area. We evaluated the implementation of objective diagnostic verification in patients with a previous diagnosis of asthma. Objective diagnostic testing might also be beneficial in individuals who are suspected of having asthma for the first time. The cost-effectiveness profile of objective testing for such a population may be different than for individuals with an existing diagnosis. Further, we did not investigate to what extent individual characteristics can be used to refine the population for whom diagnostic verification will be beneficial. For instance, performing a diagnostic workup only among patients whose asthma diagnoses were not initially made through objective verification might further improve its cost-effectiveness. We did not have access to a reliable source of evidence to infer the performance of diagnostic verification across such subgroups. However, previous objective verification testing should not necessarily preclude an objective re-evaluation of asthma, due to potentially inaccurate recall of the initial diagnosis, as well as the fact that asthma may become clinically dormant for extended periods of time, justifying treatment cessation among individuals in whom reversible airway obstruction cannot be reproduced.

In summary, our results indicate that secondary diagnostic verification of asthma during routine clinical encounters among adults with a self-reported asthma diagnosis may be cost-saving and may improve quality of life of patients. The overall conclusions made in this study about the merits of such diagnostic verification remained robust against changes in assumptions and parameter values. Nevertheless, evidence on the performance of diagnostic verification largely comes from research settings, some of which from countries other than the US. As the ‘real-world’ adherence to, and performance of, stepwise diagnostic algorithms remain untested, future implementation of such algorithms should also be coupled with subsequent evaluations.

## Data Availability

This is a simulation study so there are no underlying data. One part of the study was based on data from the Medical Expenditure Panel Survey in the US. Eligible clients can apply for such data through the relevant portal.

https://drive.google.com/file/d/1pd2e9qEa6nIMV9UpCPX4mI2SUTaUr9Yw/view?usp=sharing

## Abbreviations

ACT: Asthma Control Test
FEV_1_: Forced Expiratory Volume
GBD: Global Burden of Disease
GINA: Global Initiative for Asthma
ICER: Incremental Cost Effectiveness Ratio
ICS: Inhaled Corticosteroids
LTRA: Leukotriene receptor antagonist therapy
MCT: Methacholine Challenge Test (Bronchoprovocation Test)
OR: Odds Ratio
PC20: The concentration of methacholine needed to produce a 20% fall in FEV_1_
QALY: Quality Adjusted Life Year
US: United States

